# Demographic Factors, Attitudes, and Pre-Existing Conditions Impacting COVID-19 Vaccine Hesitancy

**DOI:** 10.1101/2025.06.30.25330574

**Authors:** Elizabeth L Kudron, Gabriela R Reguero, Katie M Marker, Jessica Guerra Callaway, Lauren A Vanderlin, Christopher R Gignoux, Colorado Center for Personalized Medicine, Jan T Lowery, Randi K Johnson

**Author notes:** **Corresponding Author** Randi K. Johnson, PhD, MPH 1890 N. Revere Court, Aurora, CO, 80045.

## Abstract

We aimed to understand factors that distinguish vaccine hesitancy from early uptake or refusal to inform public health strategies for the next emergent infectious disease pandemic and clinician-patient communication around vaccine decision-making. The study included 10,176 participants of the Colorado Center for Personalized Medicine biobank who completed a survey in Summer 2021 which included information on COVID-19 vaccine hesitancy, attitudes, and uptake. Survey data were linked to electronic health records prior to the COVID-19 pandemic to catalogue pre-existing health conditions. Early COVID-19 vaccine uptake was high (N=9,873, 96.1%), consistent with high health literacy in this population. While many demographic factors, vaccine considerations, and pre-existing conditions distinguished early uptake from hesitancy or refusal (*P*<0.05), only two characteristics distinguished the hesitant from the refusal group. Hesitant individuals were more likely than refusers to report good (31.9% versus 25.8%) or fair or poor (16.4% versus 13.4%) health. Also, they were less likely to report trust in scientists as a key factor impacting their decision to receive the COVID-19 vaccine (15.9% versus 25.8%). These findings highlight the urgent need for more tailored public health strategies to reach the distinct group of individuals who remain open to vaccination. Healthcare professionals may be especially well-positioned to address these concerns in clinical conversations by connecting individual health status and pre-existing conditions to the benefits of vaccination.

**Clinical Trials Registry:** N/A

**Highlights:**

- Many factors distinguish uptake of COVID-19 vaccines from hesitancy or refusal.
- Poorer health and trust in science differentiated hesitancy from refusal.
- These results may guide early vaccine uptake and readiness for future pandemics.

## INTRODUCTION

Widespread vaccination is one of the most effective public health strategies for mitigating the spread and severity of infectious diseases, including COVID-19. The rapid development and emergency use authorization of COVID-19 vaccines in late 2020 marked a significant milestone in the global response to the pandemic. However, despite the availability of highly effective vaccines, hesitancy to receive them emerged as a significant barrier to pandemic control^1, 2^. Vaccine hesitancy refers to a delay in acceptance or refusal of vaccines despite the availability of vaccination services^3^. Understanding the underlying factors that contribute to vaccine hesitancy is essential not only for increasing COVID-19 vaccine uptake but also for preparing for future public health crises^4,5^.

Previous research has elucidated several determinants of COVID-19 vaccine hesitancy. Sociodemographic characteristics associated with vaccine hesitancy include younger age, lower income, lower educational attainment, uninsured status, and certain racial and ethnic minority groups^6,7,8^. Attitudinal factors, including mistrust in government or pharmaceutical companies, concerns about safety and side effects, a perceived lack of urgency, and belief in natural immunity, have contributed significantly to vaccine hesitancy^6,9,10^. Comorbidities, such as obesity, diabetes, and chronic lung disease, which are typically risk factors for severe COVID-19 disease, have been shown to influence perceptions of risk and urgency, but do not uniformly drive individuals toward early vaccination^6,11^.

The literature has often categorized vaccine-hesitant individuals alongside those who firmly reject vaccines, potentially obscuring meaningful distinctions between these groups. Unlike vaccine refusers, who are unlikely to be persuaded regardless of circumstance, hesitant individuals often sit on a continuum of indecision and may be influenced by credible information, trusted messengers, or changes in personal circumstances^12^. This subgroup may harbor concerns or uncertainties that can be addressed through targeted, empathetic communication by healthcare professionals^9,10^. Differentiating between these populations is crucial, as it offers actionable insights into how to increase vaccine uptake during ongoing and future public health crises.

In this study, we investigate the characteristics and attitudes associated with early COVID-19 vaccine uptake, hesitancy, and refusal among a population enrolled in a large academic biobank. This analysis draws upon previously published survey data characterizing the broader impact of the COVID-19 pandemic and integrates electronic health record (EHR) data to provide a comprehensive profile of vaccination behaviors among a population with high health literacy^13^. By differentiating hesitant individuals from refusers, we aim to identify modifiable factors that may inform future public health strategies and clinician-patient communication around vaccine decision-making.

## MATERIALS AND METHODS

### Study Population

The Colorado Center for Personalized Medicine (CCPM) biobank was jointly launched in 2015 by the University of Colorado Anschutz Medical Campus and UCHealth as a resource to accelerate biomedical research and clinical translational efforts in the UCHealth patient population. The initiation, maintenance, and innovation of this vertically integrated genomic learning health system has been described in detail previously^14^. Current enrollment occurs electronically through a self-consent model, where patients access the study consent directly in the electronic patient portal. Any adult (18 years and older) UCHealth patient is eligible to participate. Participants provide broad consent for collection of blood, return of clinically actionable results, use of data for research, and recontact. More than 250,000 adults are enrolled in the CCPM biobank research study; this group is largely representative of the 2.5 million UCHealth patient population across Colorado and its surrounding states^14^

In response to the global COVID-19 pandemic, we surveyed all living CCPM biobank participants with a valid email address in 2020 and 2021. We published previously the survey instruments, administration procedures, and key findings around COVID-19 prevalence, testing behaviors, symptoms, and electronic health record (EHR) case ascertainment^13^. Briefly, surveys were delivered by email and study data were collected using REDCap electronic data capture tools hosted at the University of Colorado. REDCap (Research Electronic Data Capture) is a secure, HIPPA-compliant ddatabase and research management platform (Harris 2009). The resulting data was de-identified, connected to other available data elements (genotypes, EHR), and made available to researchers in a datamart engineered by Health Data Compass, the systemwide data warehouse for the University of Colorado Anschutz Medical Campus and UCHealth. Of the 180,599 eligible CCPM biobank participants enrolled as of May 31, 2021, 25,075 (13.9%) completed at least one COVID-19 survey.

We restricted this study to the 10,256 participants who answered version 2 of the survey, which included information about COVID-19 vaccine hesitancy, attitudes, and uptake^13^. Version 2 was administered in Summer 2021, after the COVID-19 vaccines became broadly distributed and available in Spring 2021. The final analysis included 10,176 participants who did not have any missing demographics or information regarding their vaccine uptake, age at dosing, or intent (**Figure 1**).

**Figure 1:**
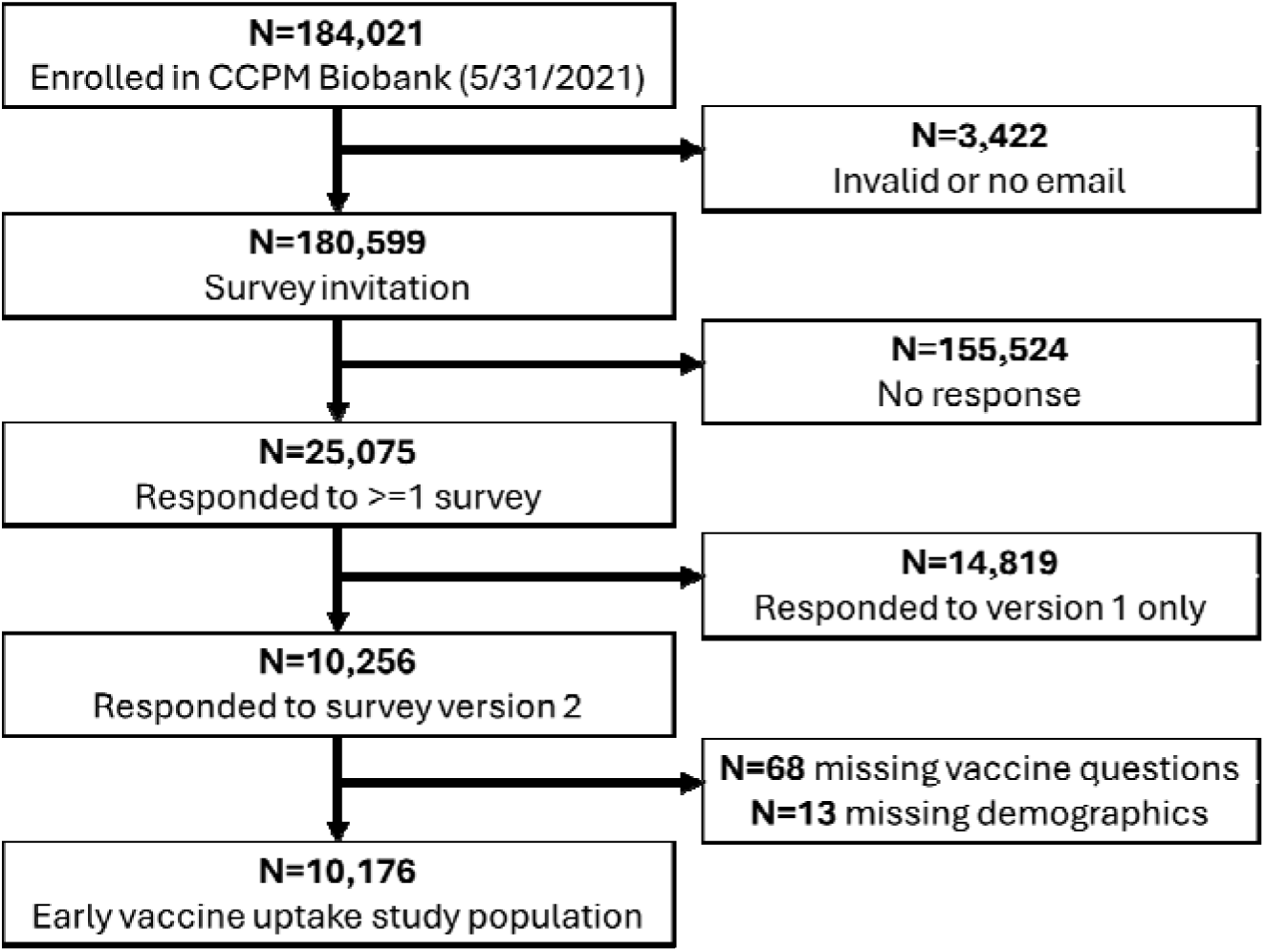
Early COVID-19 vaccine uptake study population.

### Vaccine Uptake and Factors

Respondents who reported receiving at least one dose of a COVID-19 vaccine were considered to have vaccine “uptake”. The remainder were further subdivided based on their self-reported intention to receive the vaccine. Hesitant participants reporting an intention to never receive a COVID-19 vaccine were categorized as vaccine “refusal”, and the remainder were considered “hesitant”.

All respondents were asked “What factors affect your attitudes and thoughts about getting a COVID-19 vaccine?” Respondents selected any factors that applied from a list of 11 provided options, including “Other factors”. Respondents who reported an intention to never receive a COVID-19 vaccine were asked “What are your reasons for not getting the vaccine?”

### Pre-existing Conditions and Covariates

Sociodemographic and population descriptors obtained from the EHR and the survey were categorized consistent with prior research in CCPM^13–15^ and to maintain at least 10 respondents per level for privacy. Sex, race and ethnicity, and zip code were obtained from EHR data. We categorized race and ethnicity into a single binary race-ethnicity variable with the levels Non-Hispanic White and Other for analysis. The “Other” race-ethnicity category includes Non-Hispanic Black, Hispanic, Asian, American Indian and Alaska Native, Native Hawaiian and Other Pacific Islander, and Non-Hispanic Other. We categorized respondents into young adult, mid adult, adult, and older adult categories based on the age at survey completion: 18-29, 30-44, 45-64, 65+. Survey respondents self-reported other descriptive variables, including: highest educational level obtained, current employment status, overall health status, current smoking status, and whether any household member had tested positive for COVID-19. Smoking status was defined as participants currently smoking at least one of these: cigarettes, cigars, e-cigarettes, nicotine, or marijuana.

We determined pre-existing health conditions from EHR records for survey respondents who were active in the UCHealth system prior to the COVID-19 pandemic. An active patient was defined as having at least three face-to-face encounters with the UCHealth system between January 2015 and January 2020. We used the PheWAS R package^16^ to identify algorithmic phenotypes, or phecodes, from ICD-9 and ICD-10 billing codes during the pre-pandemic window. A participant was considered to have been diagnosed with a pre-existing condition if any corresponding phecodes occurred at least three times during this time frame. **Supplemental Table 1** shows the list of phecodes associated with each pre-existing condition category. For example, an individual who had three occurrences of phecode 278 (Overweight, obesity and other hyperalimentation), 278.1 (obesity), or 278.11 (morbid obesity) in their EHR between January 2015 and January 2020 was considered to have pre-existing obesity. Additional detail on ascertainment and algorithmic phenotyping of pre-existing conditions is available in Brice et al^15^.

### Statistical Analysis

We described the distribution of sociodemographic, influencing factors, and pre-existing conditions across the uptake, hesitant, and refusal vaccine groups. Statistical differences between groups were calculated using analysis of variance test (ANOVA) for continuous variables and chi-square tests for categorical variables. Statistical significance was assessed using the overall test across the three groups (*P* for trend) at an alpha of 0.05. For any significant differences overall, we then conducted pairwise tests to identify which groups were different using adjusted chi-square tests for categorical variables and Tukey’s test for continuous variables. We conducted data analyses in R version 4.4.2.

## RESULTS

### Early COVID-19 vaccine uptake

Out of 10,176 respondents to 2021 survey, 9,783 (96.1%) had received a COVID-19 vaccination (“uptake”), 186 (1.8%) stated an intention to never receive the vaccine (“refusal”), and 207 (2.0%) had not yet been vaccinated against COVID-19 (“hesitant”). These groups significantly differed (*P* for trend<0.05) across many sociodemographic and health factors, including: sex, age, educational attainment, employment, median income based on zip code of residence, overall health status, current smoking status, and whether a household member had tested positive for COVID-19 (**Table 1**).

**Table 1:**
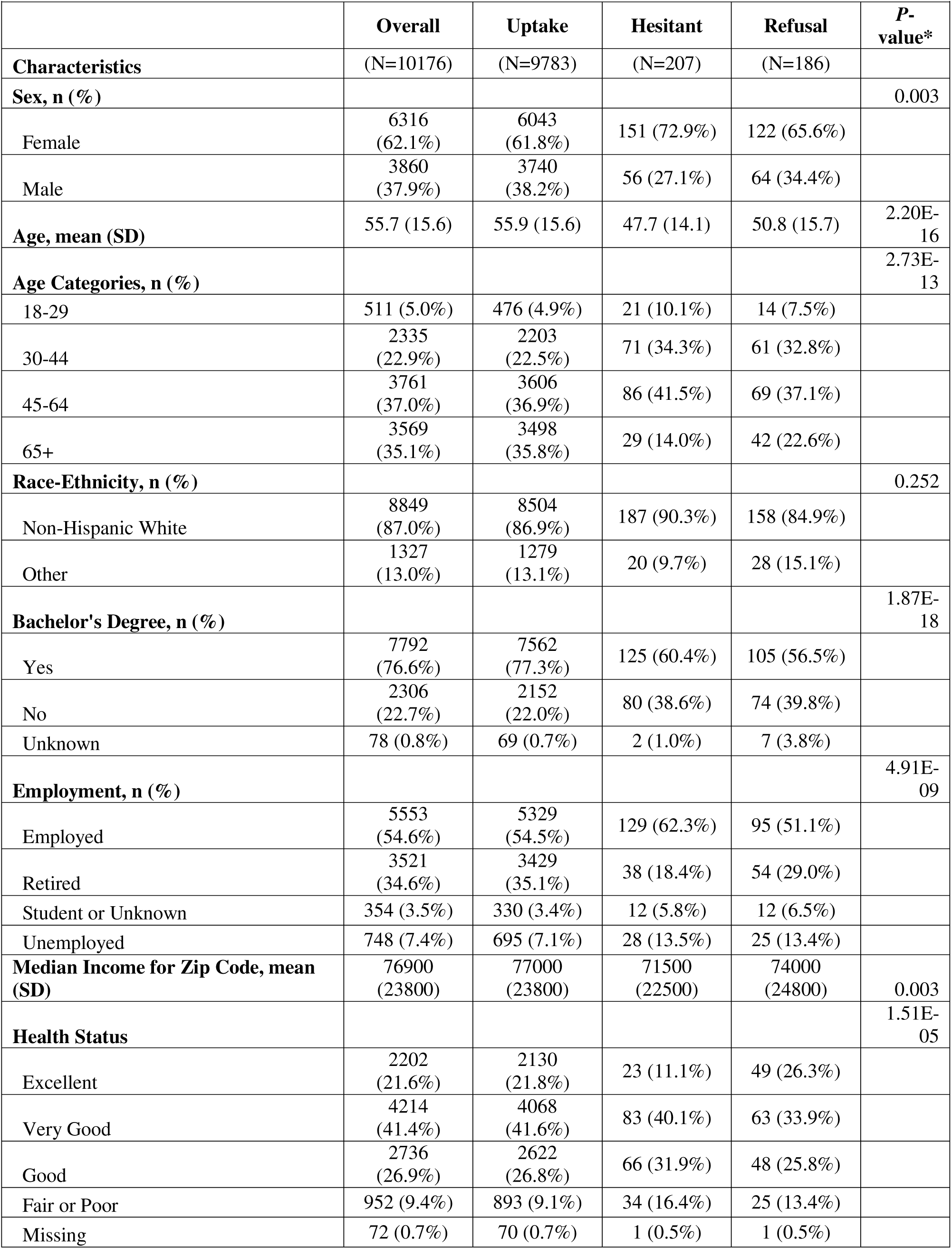
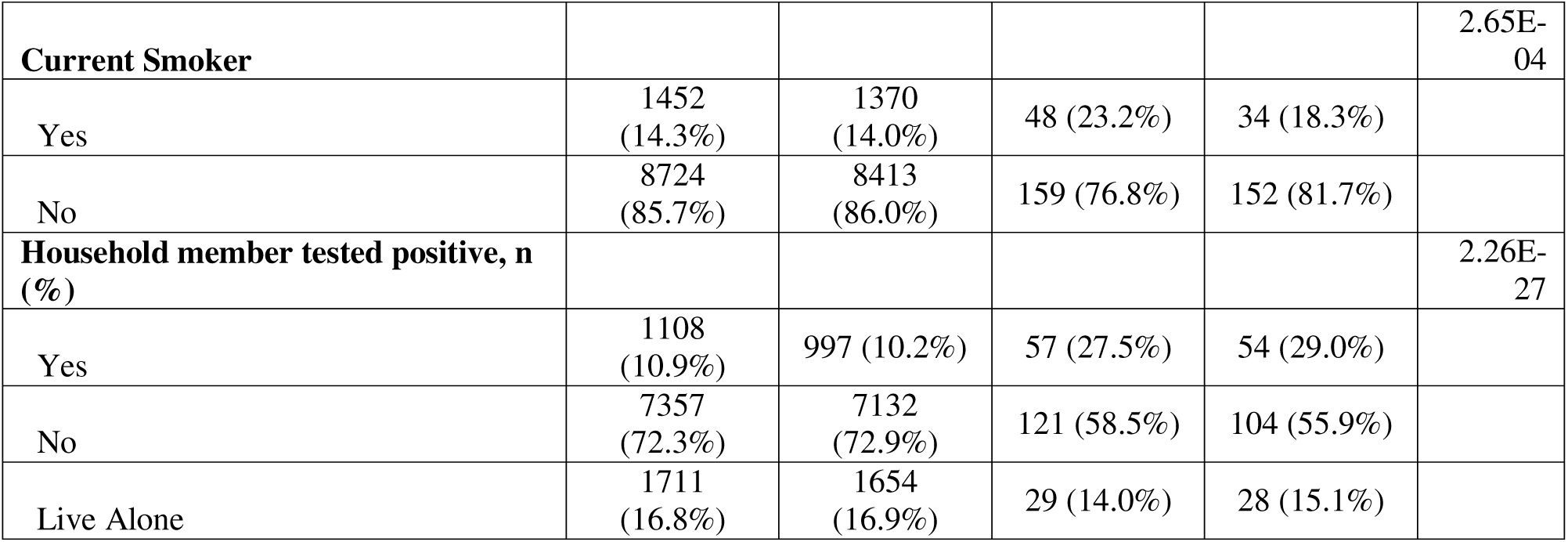
Characteristics of COVID-19 survey population by vaccine uptake status.

Pairwise comparisons indicated that the early group differed from the hesitant group and from the refusal group for most characteristics. A higher proportion of males reported early uptake (38.2%) compared to the hesitant group (27.1%). Older adults 65+ years were also more likely to have early uptake (35.8%) compared to the hesitant (14.0%) and refusal (22.6%) groups. Compared to those reporting early uptake, hesitant respondents were less likely to hold a bachelor’s degree (60.4% vs. 77.3%), had lower median income ($71,500 vs. $77,000), and were more likely to be unemployed (13.5% vs. 7.1%), a current smoker (23.2% vs. 14.0%), and have prior household exposure to COVID-19 (27.5% vs. 10.2%).

Overall, self-reported health status was the only characteristic we examined that significantly distinguished each of the three groups from each other. A higher proportion of hesitant respondents self-reported “good” (31.9%) or “fair or poor” (16.4%) health compared to both the early uptake (26.8%, 9.1% respectively) and refusal groups (25.8%, 13.4% respectively). The refusal group contained the highest proportion of participants with excellent overall health at 26.3% compared to 21.8% for early uptake and 11.1% for hesitant.

### Factors affecting early COVID-19 vaccine uptake

Eight out of 11 factors affecting thoughts and attitudes about getting a COVID-19 vaccine differed by vaccine uptake group (*P* for trend<0.05, **Figure 2**). On average, those in the unvaccinated groups (hesitant, refusal) reported more factors affecting their vaccination decision (3.49, 3.99) than those in the early uptake group (3.26). The three factors most commonly reported by the unvaccinated groups (hesitant, refusal) included the timeline in which the vaccines were developed and approved (71.5%, 67.2%), the frequently changing messages around COVID-19 (62.3%, 64.5%), and their own reading and research on coronavirus (COVID-19) vaccines (56.5%, 65.6%). A majority of the refusal group (52.2%) also reported their trust in public health officials as a key factor in their decision never to get vaccinated. In contrast, a majority of those receiving early vaccination indicated their trust in scientists (74.2%), their trust in doctors (67.0%), their trust in public health officials (55.7%), and their own reading and research on coronavirus (COVID-19) vaccines (59.5%) as key factors affecting their decision to get vaccinated. More than 950 individuals indicated “other” factors influencing their decision to get a COVID-19 vaccine, including 9.4% of the early uptake group, 29.0% of the hesitant group, and 36.0% of the refusal group.

**Figure 2:**
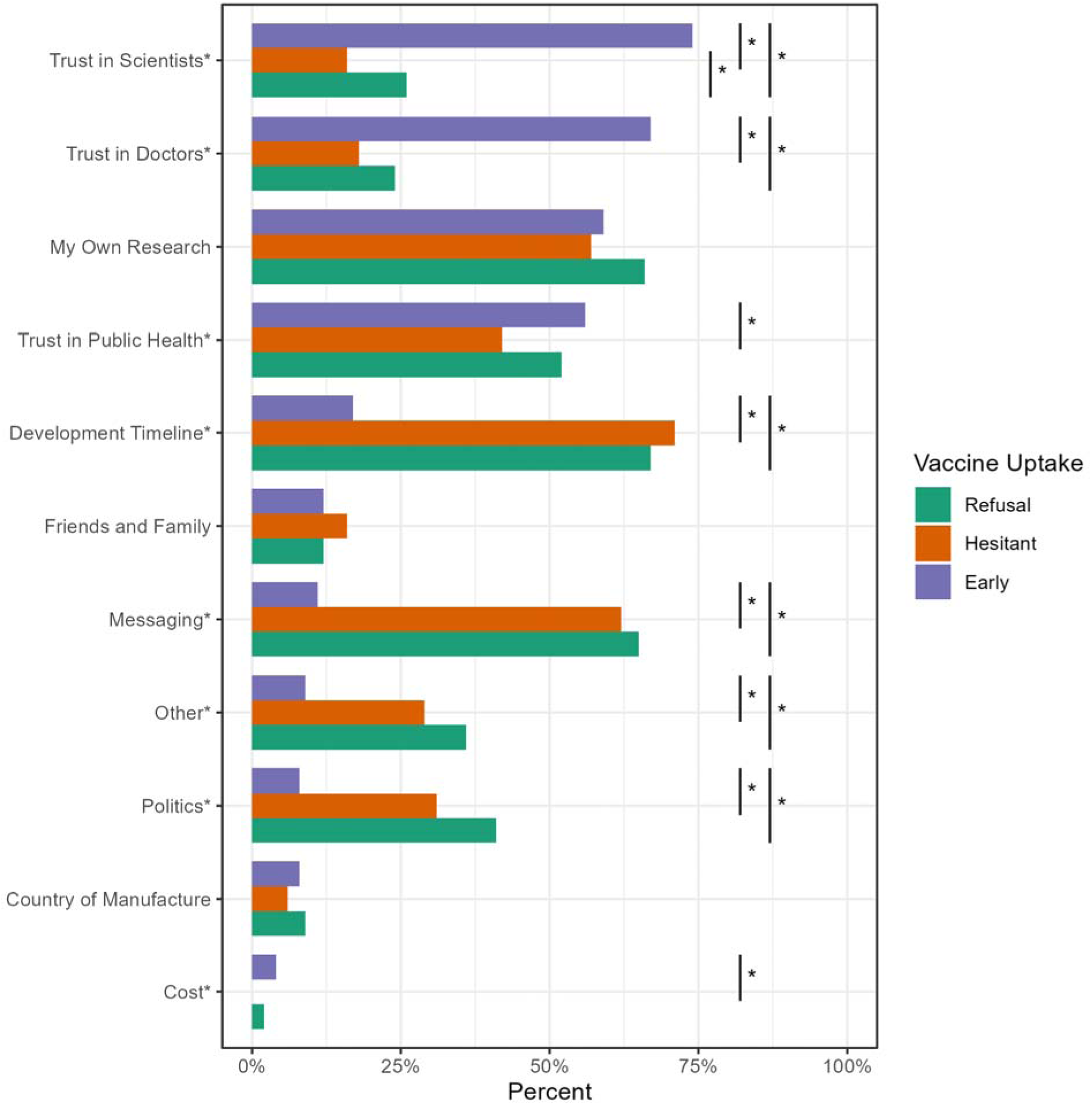
Factors influencing the decision for early, hesitant, or refusal of COVID-19 vaccination. Multiple factors could be selected. Asterisks by labels indicate significant differences (P<0.05 for trend) across the three groups. Bars with asterisks indicate significant (P<0.05) pairwise comparisons between the indicated groups.

Figure 3 summarizes the reasons given for COVID-19 vaccine refusal (N=186). The top two reasons, cited by a majority of respondents, were concerns about vaccine safety (60.8%) and side effects (55.4%). On average, respondents cited >2 reasons for the intention to never receive a COVID-19 vaccination. More than 40% cited other reasons.

**Figure 3:**
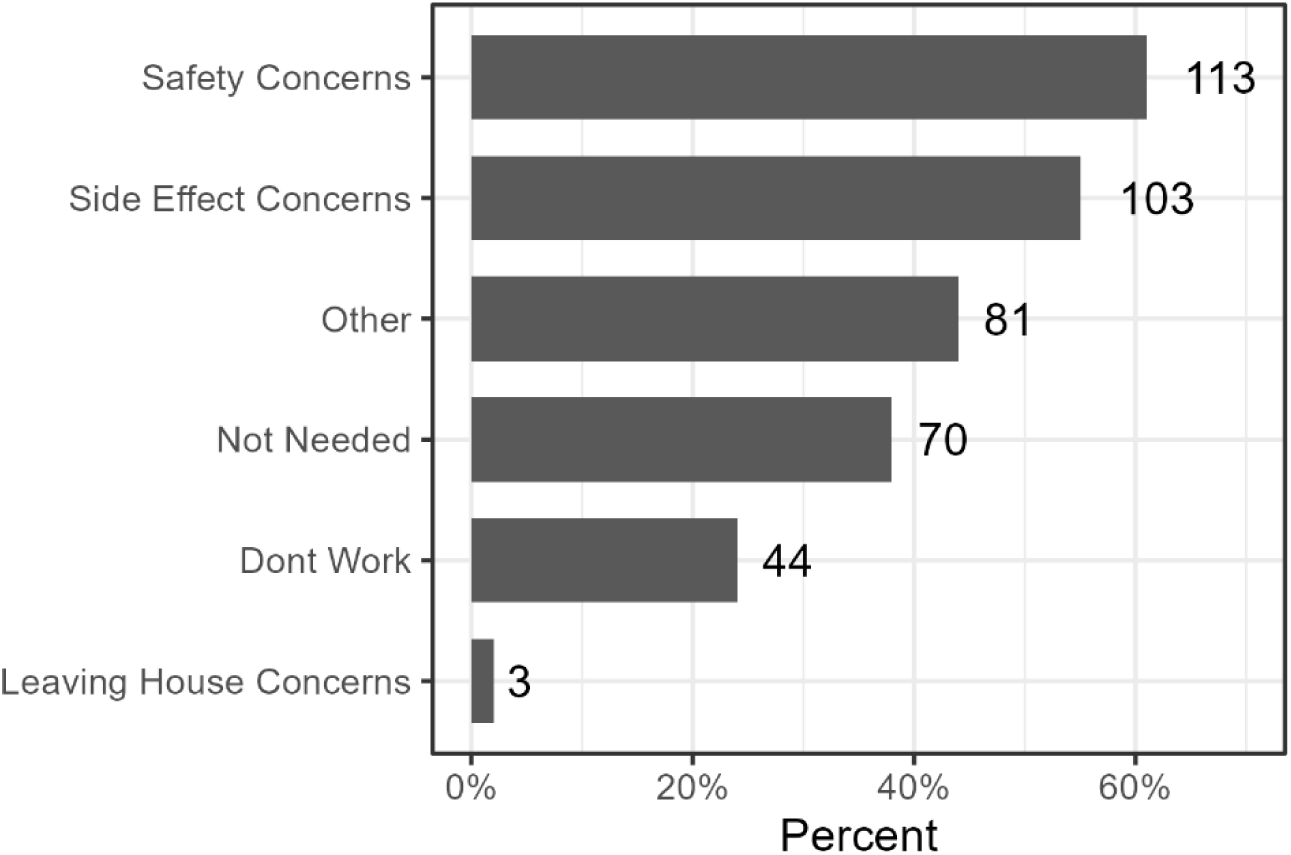
Reasons for vaccine refusal. Count and proportion of vaccine refusal respondents (N=186) reporting each indicator as a factor in their decision to never receive a COVID-19 vaccination. Multiple factors could be selected.

### Pre-existing conditions and COVID-19 vaccination uptake

Of the 10,176 survey participants, 7,915 (77.8%) met our definition of active UCHealth patients pre-COVID-19 and were included in the analysis of pre-existing conditions. This subset had comparable representation by vaccine uptake groups as the full analysis population: 7,597 (96.0%) early uptake, 161 (2.0%) hesitant, and 157 (2.0%) refusal (**Table 2**). Over 60% of this group had at least one pre-existing condition; hypertension (29.1%), heart disease (20.5%), and neoplasia (20.4%) were the three most common pre-existing conditions. Unvaccinated participants (hesitant, refusal) had higher proportions (*P* for trend<0.05) of pre-existing obesity (26.7%, 26.1%) and substance use (15.5%, 14.0%) compared to early uptake participants (obesity = 18.3%, substance use = 8.1%). There was no difference in vaccine uptake based on pre-existing diabetes, heart disease, hypertension, kidney disease, lung disease, or neoplasia. There was no significant difference in the rate of pre-existing conditions for the hesitant vs refusal groups. The average number of pre-existing conditions was similar across groups (uptake 1.31; hesitant, 1.36; refusal, 1.31).

## DISCUSSION

In this study, we examined sociodemographic factors, attitudes, and pre-existing health conditions associated with early COVID-19 vaccine uptake, hesitancy, and refusal within a population enrolled in a large academic biobank. Understanding the distinctions between individuals who are vaccine-hesitant and those who outright refuse vaccination is crucial for public health efforts aimed at promoting vaccine uptake, especially in preparing for future infectious disease outbreaks. While we found strong differences between early adopters and those who delayed or refused vaccination, surprisingly, the differences between the hesitant and refusal groups were minimal.

Consistent with prior research, we found that vaccine hesitancy was more common among females, current smokers, individuals with lower educational attainment, lower income, and those who were unemployed^6,8,17^. These findings point to persistent structural and informational inequities that affect health behavior and access. Interestingly, hesitant individuals were also more likely to report prior household exposure to COVID-19, possibly reinforcing perceived natural immunity or ambivalence due to lived experiences of illness in the home. Indeed, studies have shown that individuals with prior COVID-19 infection are less likely to receive a COVID-19 vaccine, hypothesizing that perceived immunity is impacting this decision^18,19^.

Among all unvaccinated participants, we found that concerns regarding the rapid timeline of vaccine development, changing public health messages, and their own research on COVID-19 were the most frequently cited reasons for non-uptake. Previous studies have noted similar findings^2,6,19^. While these attitudes may reflect caution rather than outright opposition, they also expose a vulnerability in public health messaging. A majority of all three groups, including early adopters, indicated that they relied on their own research to inform decision-making around COVID-19 uptake. This suggests a crucial opportunity to shape public discourse through better information dissemination. If individuals are actively seeking information, making accurate, accessible, and widely distributed educational resources is essential to counter misinformation and build confidence.

Our findings revealed few statistically significant differences between the hesitant and refusal groups, suggesting these populations may share similar foundational concerns and characteristics. Notably, hesitant participants were more likely to report poorer self-perceived health status, whereas both early adopters and vaccine refusers were more likely to report excellent health. These findings highlight the nuanced relationship between self-rated health and vaccine uptake. While some studies have shown that self-reported poor health is associated with lower COVID-19 vaccine adoption^20^, others have noted that high satisfaction with health is associated with vaccine hesitancy^21^.

These differences may reflect divergent perceptions of personal vulnerability or vaccine efficacy. Hesitant individuals may have been more cautious or uncertain about how the vaccine would affect them personally, particularly in the early phases of the rollout when safety data were still emerging^2,19^. Conversely, hesitant individuals’ perceptions of their health as fair or poor may have exacerbated their fears of a new vaccine worsening their pre-existing conditions.

Although we found no significant differences in the overall number of pre-existing conditions across early adopters, hesitant individuals, and refusers, specific conditions varied meaningfully by group. Notably, unvaccinated participants, both hesitant and refusing, had higher rates of obesity and substance use compared to early adopters. These conditions are well-established risk factors for severe COVID-19 outcomes^22,23^, making their higher prevalence among the unvaccinated particularly concerning.

When considered alongside self-rated health, these findings offer a potential avenue for targeted communication to improve vaccine uptake. Healthcare professionals may be able to leverage nuances in perceived vulnerability during patient discussions. Hesitant individuals, who were more likely than refusers to report fair or poor health, may be more receptive to vaccine messaging that emphasizes protection from severe outcomes. In contrast, refusers were more likely to report excellent health, potentially reinforcing a sense of invulnerability. These distinctions highlight the importance of addressing perceived health risks and underlying medical conditions during clinical conversations with hesitant individuals, who may be more open to engagement than those firmly opposed to vaccination.

Trust emerged as a key differentiating factor across vaccine uptake groups. Like previous studies, early adopters were more likely to cite trust in scientists, doctors, and public health officials as a factor in vaccine uptake compared to unvaccinated individuals^6,19^. We found that hesitant participants were less likely than both vaccine adopters and refusers to report trust in scientists as a key factor impacting their decision to receive the COVID-19 vaccine. This is an important distinction as science-based arguments may be less likely to persuade hesitant individuals to vaccinate.

A limitation of our study is that we did not ask hesitant individuals about their concerns related to vaccine safety and side effects. However, these concerns were commonly cited by refusers and have been widely reported in previous literature^6,19^. Future studies would benefit from exploring the specific concerns of hesitant individuals to better delineate their position along the continuum of vaccine acceptance.

The biobank setting provided a unique and timely opportunity to collect high-quality survey and EHR-linked data in response to a rapidly evolving public health emergency. The ability to deploy a targeted survey shortly after the COVID-19 vaccines became widely available, combined with retrospective access to participants’ clinical data, allowed for novel analyses of pre-existing conditions and their associations with vaccine behavior. While biobank participants represent a somewhat self-selected and health-engaged population, the observed variability in vaccination decisions suggests that even within research-savvy groups, vaccine refusal and hesitancy persist.

Taken together, our findings highlight the urgent need for more tailored public health strategies. There is a group of individuals who remain open to vaccination but are waiting for more information or reassurance. These individuals differ from early adopters in nearly all sociodemographic and attitudinal factors we measured, but they are also distinct from those who have fully rejected vaccination. Their attitudes, marked by poorer self-perceived health and a lower reliance on trust in scientists, suggest an important target for intervention. Healthcare professionals may be especially well-positioned to address these concerns in clinical conversations by connecting individual health status and pre-existing conditions to the benefits of vaccination.

In conclusion, understanding the differences between early vaccine adopters, the hesitant, and the refusal groups is critical to improving vaccine uptake and preventing mortality and morbidity in a future pandemic. While the line between hesitancy and refusal may be thin, it represents a meaningful opportunity for engagement. Our findings provide guidance for future outreach efforts aimed at strengthening vaccine confidence and improving readiness for the next emerging infectious disease threat.

## CRediT Author Statement

**Elizabeth L Kudron**: Conceptualization, Supervision, Data Curation, Writing – Original Draft, **Gabriela R Reguero**: Formal analysis, Writing - Review & Editing, **Katie M Marker**: Data Curation, Writing - Review & Editing, **Jessica Guerra Callaway**: Formal analysis, Writing - Review & Editing, **Lauren A Vanderlin**: Methodology, Software, Data Curation, Writing - Review & Editing, **Christopher R Gignoux**: Funding acquisition, Writing - Review & Editing, **CCPM Biobank**: Resources, Funding acquisition, Data Curation, **Jan T Lowery**: Conceptualization, Project administration, Methodology, Writing - Review & Editing. **Randi K Johnson**: Conceptualization, Supervision, Formal analysis, Visualization, Writing – Original Draft

## Acknowledgements

We would like to thank all participants in the CCPM, as well as the entire CCPM team, whose dedication to contributing meaningfully to the COVID-19 emergency enabled this research.

## Data Availability

The dataset used for this study is not publicly available. Interested parties may request access to CCPM biobank data through the Access to Biobank Committee (https://medschool.cuanschutz.edu/cobiobank/investigators-industry-partners/access-to-biobank-committee-(abc)).

---

**Table 3:** Early COVID-19 vaccine uptake by pre-existing condition*.

| *Table 3: Early COVID-19 vaccine uptake by pre-existing condition\** |  |  |  |  |  |
| --- | --- | --- | --- | --- | --- |
|  | **Overall** | **Uptake** | **Hesitant** | **Refusal** | **P-value** |
| **n (%)** | (N=7915) | (N=7597) | (N=161) | (N=157) |  |
| Diabetes | 756 (9.6%) | 720 (9.5%) | 20 (12.4%) | 16 (10.2%) | 4.36E-01 |
| Heart disease | 1621 (20.5%) | 1554 (20.5%) | 35 (21.7%) | 32 (20.4%) | 9.23E-01 |
| Hypertension | 2303 (29.1%) | 2225 (29.3%) | 36 (22.4%) | 42 (26.8%) | 1.29E-01 |
| Kidney disease | 421 (5.3%) | 406 (5.3%) | 9 (5.6%) | 6 (3.8%) | 6.93E-01 |
| Lung disease | 1050 (13.3%) | 995 (13.1%) | 30 (18.6%) | 25 (15.9%) | 7.49E-02 |
| Neoplasia | 1615 (20.4%) | 1558 (20.5%) | 30 (18.6%) | 27 (17.2%) | 5.08E-01 |
| Obesity | 1471 (18.6%) | 1387 (18.3%) | 43 (26.7%) | 41 (26.1%) | 1.20E-03 |
| Substance use | 665 (8.4%) | 618 (8.1%) | 25 (15.5%) | 22 (14.0%) | 1.40E-04 |
| At least one pre-existing condition | 4797 (60.6%) | 4589 (60.4%) | 104 (64.6%) | 104 (66.2%) | 1.93E-01 |
| Number of pre-existing conditions, mean (SD) | 1.20 (1.31) | 1.19 (1.31) | 1.36 (1.35) | 1.31 (1.30) | 1.61E-01 |
| *Pre-existing conditions were defined by the presence of at least three occurrences of a phecode in a participant's EHR between January 2015 and January 2020. Phecodes included for each condition are listed in Supplemental Table 1. |  |  |  |  |  |

**Supplemental Table 1:** Map of phecodes used to define pre-existing conditions from electronic health records in the Colorado Center for Personalized Medicine biobank.

| **Pre-existing condition** | **Phecodes** |
| --- | --- |
| Diabetes | 249, 250, 250.1, 250.11, 250.12, 250.13, 250.14, 250.15, 250.2, 250.21, 250.22, 250.23, 250.24, 250.25, 250.3, 250.6, 250.7 |
| Heart Disease | 394, 394.1, 394.2, 394.3, 394.4, 394.7, 395, 395.1, 395.2, 395.3, 395.4, 395.6, 411, 411.1, 411.2, 411.4, 411.41, 411.8, 411.9, 414, 414.2, 426, 426.2, 426.21, 426.22, 426.23, 426.24, 426.25, 426.3, 426.31, 426.32, 426.4, 426.8, 426.9, 426.91, 426.92, 427, 427.1, 427.11, 427.12, 427.2, 427.21, 427.22, 427.3, 427.4, 427.41, 427.42, 427.5, 427.6, 427.61, 428, 428.1, 428.2, 428.3, 428.4, 429, 429.1 |
| Hypertension | 401, 401.1, 401.2, 401.21, 401.22, 401.3 |
| Kidney Disease | 585, 585.1, 585.2, 585.3, 585.31, 585.32, 585.33, 585.34, 585.4, 587 |
| Lung Disease | 495, 495.1, 495.11, 495.2, 496, 496.1, 496.2, 496.21, 499, 508, 509, 509.1, 509.2, 509.3, 509.5, 510.2 |
| Neoplasia | 145, 145.1, 145.2, 145.3, 145.4, 145.5, 149, 149.1, 149.2, 149.3, 149.4, 149.5, 149.9, 150, 151, 153, 153.2, 153.3, 155, 155.1, 157, 158, 159, 159.2, 159.3, 159.4, 164, 165, 165.1, 170, 170.1, 170.2, 172, 172.1, 172.11, 172.2, 172.21, 172.22, 172.3, 173, 174, 174.1, 174.11, 174.2, 174.3, 180, 180.1, 180.3, 182, 184, 184.1, 184.11, 184.2, 185, 187, 187.1, 187.2, 187.8, 189, 189.1, 189.11, 189.12, 189.2, 189.21, 189.4, 190, 191, 191.1, 191.11, 193, 194, 195, 195.1, 195.3, 198, 198.1, 198.2, 198.3, 198.4, 198.5, 198.6, 198.7, 199, 199.4, 200, 201, 202, 202.2, 202.21, 202.22, 202.23, 202.24, 204, 204.1, 204.11, 204.12, 204.2, 204.21, 204.22, 204.3, 204.4, 209, 230 |
| Obesity | 278, 278.1, 278.11 |
| Substance Use | 316, 316.1, 317, 317.1, 317.11, 318 |

